# An outbreak of Salmonella Typhimurium following an Eid al-Adha celebration barbecue in Wales (UK), July 2021

**DOI:** 10.1101/2023.01.03.22283848

**Authors:** James Adamson, Clare Sawyer, Gemma Hobson, Emily Clark, Laia Fina, Oghogho Orife, Robert Smith, Christopher Williams, Harriet Hughes, Allyson Jones, Sarah Swaysland, Oluwaseun Somoye, Ryan Phillips, Junaid Iqbal, Israa Mohammed, George Karani, Daniel Rh Thomas, the Incident Management Team

**Author notes:** **Corresponding author** James Adamson, CDSC Public Health Wales, Cardiff, Wales, UK, [0044] (0)29 2022 7744. Joint first authors (James Adamson, Clare Sawyer).

## Abstract

**Background:** On a Friday evening in July 2021, Public Health Wales received notification of two cases of salmonella gastroenteritis. Both cases reported attending an Eid al-Adha celebration barbecue in a public park in Cardiff, UK two days earlier. Case finding over the weekend indicated further cases in those who had attended this event and an outbreak investigation was initiated.

**Methods:** Cases were defined as an individual with diarrhoea and/or vomiting with date of onset on the day of the barbecue, with an epidemiological link to the Eid-al Adha celebration meal. We undertook a cohort study in 36 people attending the barbecue and an associated lunchtime event, and a nested case-control study using Firth logistic regression. A communication campaign that was sensitive towards cultural practices was developed in collaboration with the community, community liaison staff and the Public Health Wales communications team.

**Results:** Consumption of a traditional raw liver dish, ‘marrara’, at the barbecue was the most likely vehicle for infection (Firth logistic regression, aOR: 49.99, 95%CI 1.71-1461.54, p=0.02). Meat came from two local butchers with the same supplier and food samples yielded identical whole genome sequences to those from cases. This outbreak highlights the need for appropriate food hygiene advice in communities preparing traditional dishes.

**Conclusions:** This outbreak identified a new vehicle of interest, a traditional raw offal dish. Future outbreak investigations, particularly where cultural events are associated with particular foods, should consider dishes beyond those on routine questionnaires to those which may be relevant to the community in question.

## Introduction

*Salmonella enterica* is a zoonotic, bacterial pathogen which is a common cause of gastrointestinal disease in humans [1], which is usually self-limiting. The most commonly reported serotypes of *Salmonella enterica* in the UK are *Salmonella Enteritidis* followed by *Salmonella Typhimurium* [2]. These two serotypes account for half of all salmonella laboratory reports in England and Wales [3]. Transmission of salmonella typically occurs through consumption of contaminated food or water, or through contact with animals and their environments [4]. Contamination of food and produce with salmonella can occur at any point as a result of poor hygienic practices during the production, storage or preparation of food. In the UK, an estimated 39,000 cases of non-typhoidal salmonellosis occur in the community per year, of which, around 10,000 are notified [5].

Foodborne outbreaks caused by salmonella in the UK are usually attributed to poultry and poultry products [6]. As a result, salmonella controls in livestock in the UK are targeted at poultry, and data on salmonella in poultry are collected as part of statutory surveillance systems [7]. Most Salmonella reports from cattle, sheep and pigs arise following investigation of diseased animals.

On a Friday in July 2021, Public Health Wales (PHW) Microbiology reported two cases of gastrointestinal disease with a positive PCR result for salmonella. Follow up by Public Health Wales and Regulatory Services for Bridgend, Cardiff & Vale of Glamorgan Councils established that both cases were in individuals from the same Cardiff North African community who reported attending a barbecue in a public park two days earlier to celebrate Eid al-Adha. Celebration lunches had also been held on the same day. We initiated an epidemiological investigation in order to identify the source and extent of the outbreak, and to implement control measures to prevent further spread.

## Methods

### Case finding

Initial case finding was carried out by the Public Health Wales out-of-hours health protection service, in collaboration with environmental health and Public Health Microbiology. Cases were defined as shown in table 1.

**Table 1.** Case definitions used during the outbreak investigation.

| Case definition | Rationale |
| --- | --- |
| Possible outbreak case | Gastroenteritis case (diarrhoea and/or vomiting) with date of onset on day of barbecue, WITH an epidemiological link (i.e. resident in same household as attendee) to the Eid-al Adha celebration barbecue or lunch |
| Probable outbreak case | Meets criteria for possible case AND has a PCR positive result for *Salmonella* Typhimurium with or without a positive culture result<br>**OR**<br>Gastroenteritis case (diarrhoea and/or vomiting) with date of onset on day of barbecue, AND attended the Eid-al Adha celebration barbecue or lunch |
| Confirmed outbreak case | Meets criteria for a probable case and has a culture positive result for *Salmonella* Typhimurium |
| Cohort study case | An individual who attended the Eid-al Adha barbecue, who became unwell with diarrhoea and/or vomiting within 8 hours - 72 hours following the event and for whom a completed questionnaire was available |
| Whole genome sequenced case | An individual with *Salmonella* Typhimurium t5.4574.%(1.26.136.634.2037.4574.%) |

All cases identified were interviewed and risk of onward transmission was assessed. Standard advice was given to cases and their contacts about infection prevention and control, and where necessary, cases were excluded from work or school. Information was collected about the personal characteristics of cases, their symptoms, food and travel history, and a line list was maintained. All cases were asked to provide a stool sample for microbiological analysis.

### Cohort study

Those attending the barbecue or a celebration lunch were included in a cohort study. The names and household residence of attendees was cross-referenced and verified by discussing who was present with cases and their household. We tested the hypothesis that being a case was associated with consumption of sheep meat products served at the barbecue and/or celebration lunch. We designed and administered a questionnaire survey to all those attending a celebration lunch and/or the evening barbecue. This questionnaire was administered by face-to-face field interviews (as the preferred option), telephone interview (second preference) or by the online questionnaire tool ‘SmartSurvey’ [8]. The majority of interviews were conducted in English, but for one telephone interview, an Arabic interpreter was used by utilising the Language Line service [9]. We asked about specific dishes consumed and the portion size of each. These details were checked with those who prepared the dishes and using online recipes. Information on demographics, clinical features, outcomes and exposures were collected, as was travel history, contact with animals and contact with infectious individuals in the 48 hours prior to the celebration lunch and barbecue events.

### Statistical analysis

We described the characteristics of cases and their exposure history, and constructed an epidemic curve. Using our cohort of people attending the barbecue and/or celebration lunch, we compared likelihood of being a case in those consuming specific food items. All analysis was undertaken using STATA 15.

We examined whether those who ate more of an implicated food were more likely to have severe illness. This dose-response analysis was conducted by asking survey respondents how much of each food they ate (a small, normal or large portion). Severity of disease was assess by using their responses about seeking medical care. Those attending hospital were considered more ill than those who did not (no cases contacted their GP) and those who were admitted to hospital were considered most ill of all.

In order to allow for zero-cell counts or unanimous consumption of certain food types, and in particular examine lamb consumption further, we conducted a Firth logistic regression using a case-control design. Significant odds ratios (P<0.05) calculated in the univariate results were included in the Firth regression.

### Microbiology and whole genome sequencing of human samples

Stool samples were sent to Public Health Wales Microbiology Cardiff for PCR testing. Those returning a positive PCR result for *Salmonella Spp* were cultured and subtyped locally. Samples of positive cultures were also sent to the UK reference laboratory (Gastro Bacterial Reference Unit (GBRU) in Colindale, UK Health Security Agency (UKHSA)) for whole genome sequencing (WGS). Full SNP addresses are reported by this service, with the 5 SNP level used for cluster designation and public health action.

### Food microbiology

Households containing a case were asked to provide samples of any leftover food products from the barbecue or lunches. Environmental and food samples were taken from the premises, local butchers and supplying abattoir. Samples were sent initially to Public Health Wales’s Food, Water and Environment (FEW) laboratories (University Hospital Llandough, Cardiff) for detection, enumeration and serotyping of Salmonella, before referring isolates to GBRU for whole genome sequencing.

## Results

### The outbreak

In total, 22 cases were identified in this outbreak, all of whom were symptomatic. All cases were part of a social network (n=43; attack rate = 51%), the majority of whom attended the barbecue (22 cases in 32 attendees) or celebration lunch. The epidemic curve (figure 2) is suggestive of a point source, with the majority of cases reporting symptom onset in the day following the barbecue (n=14), with two further cases developing symptoms the day after this. A further five cases developed symptoms in the following eight days, which is suggestive of person-to-person transmission.

The discussion with cases and their households about celebration event attendance also gave valuable insight into the wider community network and those visiting specifically for these events. We received 36 completed surveys, of which 19 were male respondents (figure 1a). Of these survey respondents, 20 individuals were cases (12 male and 8 female, figure 1b). There were no cases in the 0-9 years age group but men were affected in all other age groups, notably the 30-49 years strata. Females had no cases in all but two strata, but the female 10-19 years age group was the worst affected strata of all respondents. All survey respondents identified were of non-white ethnicity and 30 identified a family connection to the same North African country.

**Figure 1a.**
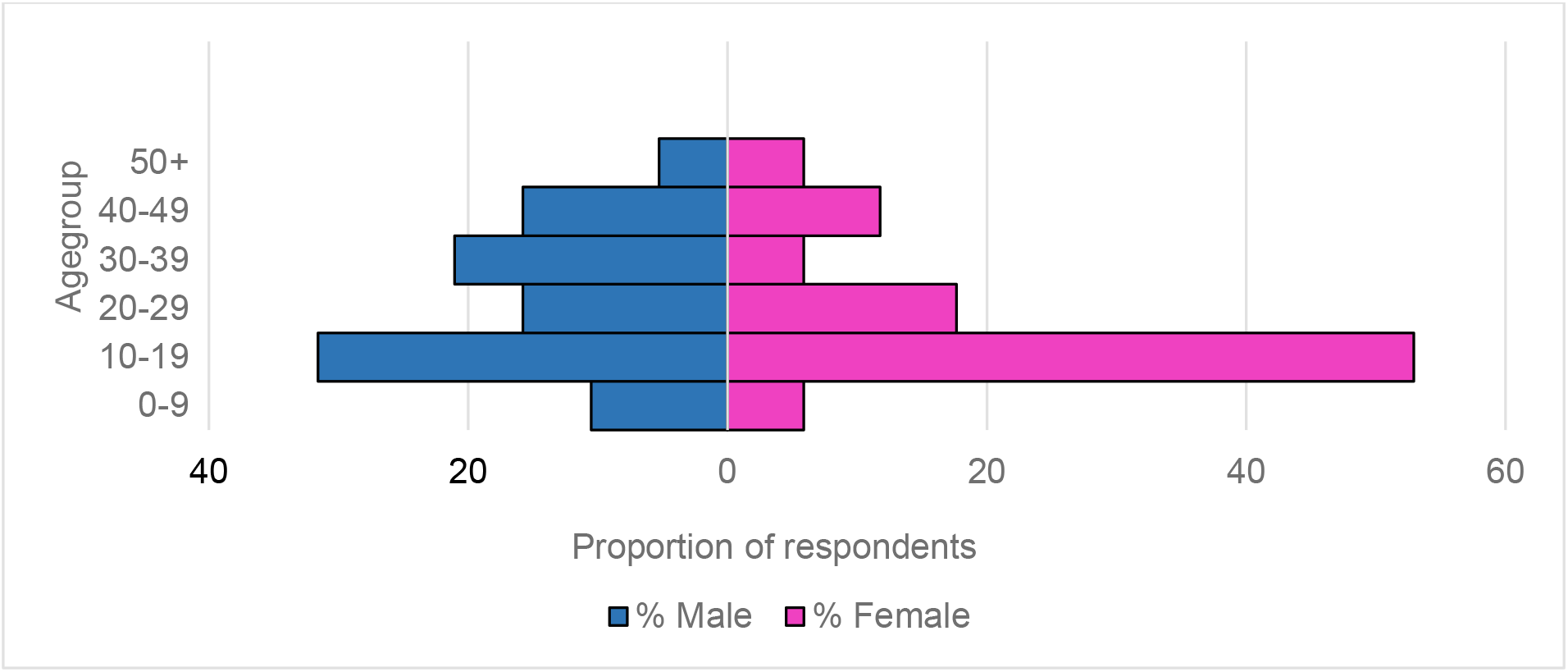
Age-sex profile of survey respondents, n=36.

**Figure 1b.**
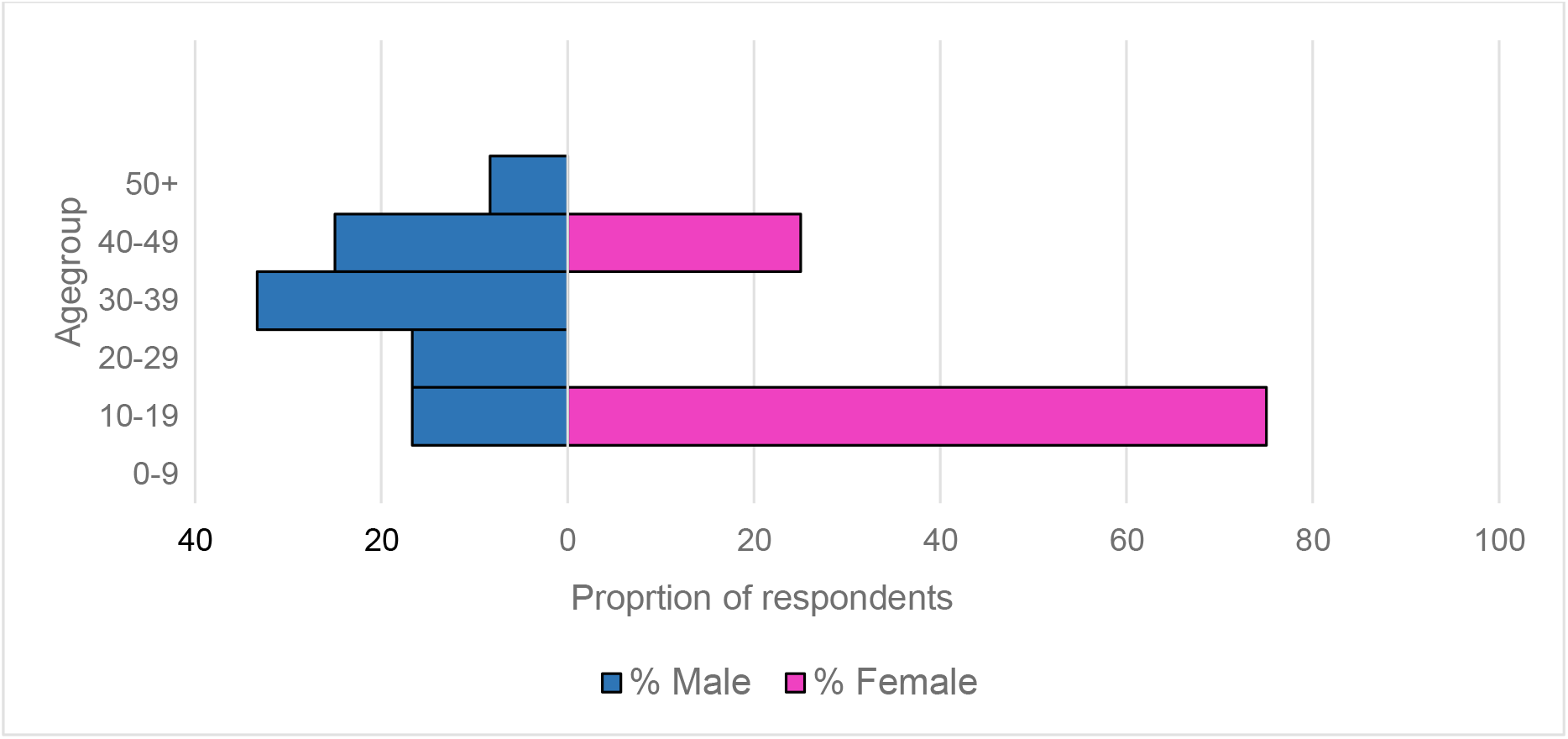
Age-sex profile of survey respondent cases, n=20.

### Cohort study

Of the 36 surveys completed, 31 lived in Cardiff, and five were visiting Cardiff (from other regions of the UK). These latter five individuals were staying with friends and family in Cardiff for the Eid celebrations and cited this trip as the only stay outside home in the three days before the barbecue. Only one other respondent mentioned travel in the three days before the barbecue, where they had visited a local beach with friends for another barbecue. Of all survey responses, 29 attended the barbecue (cases=17) and 27 attended a lunch event (cases=16). There were seven people attending only a lunch event (cases=3) and nine people attending only the barbecue (cases=4). We defined our cohort as those attending the barbecue (n=32), giving a survey response rate of 91%, but also examined what these individuals ate at lunch where applicable. There were 19 male respondents (cases=12) and 16 female respondents (cases=8). Twelve respondents reported attending emergency health services (A&E) because of their symptoms, six of these were admitted and four were kept overnight. One patient required intensive care (ITU) to manage an acute electrolyte disturbance and renal function. The two initial cases reported were part of a group from the same Cardiff North African community who had purchased jointed whole lamb carcasses and offal from two local butchers. This was prepared at home to consume together at an evening barbecue, along with a number of traditional dishes. These included “marara”, made from raw liver, usually cut into strips, washed in lemon juice and vinegar and marinated in herbs, spices and peanut butter. One respondent noted GI sickness in a barbecue attendee, resident in their household, with symptoms starting before the barbecue.

#### Univariate analysis

Univariate analysis showed an association between consumption of raw liver at the barbecue on 20 July 2021 and being a case (RR = 2.5, 95% CI: 1.46 – 4.28, p<0.05). Because all cases attending the barbecue reported consuming barbecued lamb and no non-cases ate the lamb, it was not possible to calculate a risk ratio for lamb consumption in univariate analysis. There was some evidence for an association between consumption of raw kidney and raw liver at lunchtime and being a case (RR = 1.8, 95% CI: 1.28 – 2.52, p = 0.5), although the p-value was above the cut off for significance. The full list of foods consumed at the lunch and barbecue events, and their associated risk ratios from the univariate analysis, are detailed in table 2. In the univariate analysis, there were no foods consumed at lunchtime that were significantly associated with being a case.

**Table 2.** Cohort study univariate results, barbeque and/or lunch attendees (n=29), July 2021.

| Exposure | Exposed |  |  | Unexposed |  |  | Risk Ratio | 95% (CI) | P-value |
| --- | --- | --- | --- | --- | --- | --- | --- | --- | --- |
|  | N | Cases | AR<sup>1</sup>% | N | Cases | AR% |  |  |  |
| Raw liver (BBQ) | 9 | 9 | 100 | 20 | 8 | 40 | 2.5 | **[1.46 - 4.28]** | **0.00** |
| Lamb (BBQ)<sup>2</sup> | 26 | 17 | 65 | 3 | 0 | 0 | - | **[.-]** | **0.06** |
| Pepsi (BBQ) | 9 | 3 | 33 | 20 | 14 | 70 | 0.48 | [0.18 - 1.25] | 0.11 |
| Lemonade with yoghurt (BBQ) | 10 | 8 | 80 | 19 | 9 | 47 | 1.69 | [0.96 - 2.98] | 0.13 |
| Watermelon (BBQ) | 16 | 7 | 44 | 13 | 10 | 77 | 0.57 | [0.30 - 1.07] | 0.13 |
| Stew (BBQ) | 8 | 3 | 38 | 21 | 14 | 67 | 0.56 | [0.22 - 1.45] | 0.22 |
| Kisra (bread) (BBQ) | 8 | 3 | 38 | 21 | 14 | 67 | 0.56 | [0.22 - 1.45] | 0.22 |
| Fresh tea (BBQ) | 3 | 3 | 100 | 26 | 14 | 54 | 1.86 | [1.30 - 2.65] | 0.25 |
| Cooked liver dish (BBQ) | 3 | 3 | 100 | 26 | 14 | 54 | 1.86 | [1.30 - 2.65] | 0.25 |
| Lamb (Lunch) | 12 | 9 | 75 | 17 | 8 | 47 | 1.59 | [0.87 - 2.91] | 0.25 |
| Kunafa (pastry dessert) (BBQ) | 15 | 7 | 47 | 14 | 10 | 71 | 0.65 | [0.35 - 1.23] | 0.26 |
| Lemonade alone (BBQ) | 5 | 4 | 80 | 24 | 13 | 54 | 1.48 | [0.83 - 2.62] | 0.37 |
| Mixed green salad (BBQ) | 14 | 7 | 50 | 15 | 10 | 67 | 0.75 | [0.40 - 1.41] | 0.46 |
| Raw kidney (Lunch) | 2 | 2 | 100 | 27 | 15 | 56 | 1.8 | [1.28 - 2.52] | 0.50 |
| Raw liver (Lunch) | 2 | 2 | 100 | 27 | 15 | 56 | 1.8 | [1.28 - 2.52] | 0.50 |
| Carrot salad (Lunch) | 4 | 3 | 75 | 25 | 14 | 56 | 1.34 | [0.69 - 2.60] | 0.62 |
| Other drink (BBQ) | 12 | 8 | 67 | 17 | 9 | 53 | 1.26 | [0.69 - 2.30] | 0.70 |
| Spicy sauce (BBQ) | 14 | 9 | 64 | 15 | 8 | 53 | 1.21 | [0.65 - 2.23] | 0.71 |
| Mixed carrot salad (BBQ) | 3 | 2 | 67 | 26 | 15 | 58 | 1.16 | [0.49 - 2.75] | 1.00 |
| Other meat (Lunch) | 4 | 2 | 50 | 25 | 15 | 60 | 0.83 | [0.30 - 2.34] | 1.00 |
| Pickled salad (BBQ) | 4 | 2 | 50 | 25 | 15 | 60 | 0.83 | [0.30 - 2.34] | 1.00 |
| Hummus (BBQ) | 4 | 2 | 50 | 25 | 15 | 60 | 0.83 | [0.30 - 2.34] | 1.00 |
| Basbousa (BBQ) | 10 | 6 | 60 | 19 | 11 | 58 | 1.04 | [0.55 - 1.96] | 1.00 |
| Aubergine salad (BBQ) | 7 | 4 | 57 | 22 | 13 | 59 | 0.97 | [0.47 - 2.01] | 1.00 |

<sup>1</sup>AR – Attack rate
<sup>2</sup>Risk ratio not calculable due to no unexposed cases

### Firth regression results

Further analysis was carried out using Firth regression to investigate whether lamb or raw offal consumption was important. Firth regression is based on case-control design and we used the same 29 barbecue and/or lunch attendees for our analysis.

Again, due to the fact that no controls ate raw liver and that all cases ate the lamb, univariate odds ratios could not be calculated, but a significant association was found (table 3). Firth regression was used to analyse these two variables of interest. We also examined the effect of the consumption of spicy sauce in our model because field reports had indicated that some people might have interpreted this to mean dipping cooked lamb into the marinade juices from the raw liver dish.

**Table 3.** Case-control study univariate results, barbeque attendees (n=29), July 2021.

| Exposure | Cases |  |  | Controls |  |  | Odds ratio | 95%CI | p-value |
| --- | --- | --- | --- | --- | --- | --- | --- | --- | --- |
|  | Total | Exposed | % | Total | Exposed | % |  |  |  |
| Raw liver | 17 | 9 | 52.94 | 12 | 0 | 0 | . | **[3.01-.]** | **0.002** |
| Lamb | 17 | 17 | 100 | 12 | 9 | 75 | . | **[1.27-.]** | **0.029** |
| Pepsi | 17 | 3 | 17.65 | 12 | 6 | 50 | 0.21 | [0.03-1.5] | 0.064 |
| Watermelon | 17 | 7 | 41.18 | 12 | 9 | 75 | 0.23 | [0.03-1.5] | 0.071 |
| Lemonade-yoghurt drink | 17 | 8 | 47.06 | 12 | 2 | 16.67 | 4.44 | [0.61-51.2] | 0.09 |
| Tea | 17 | 3 | 17.65 | 12 | 0 | 0 | . | [0.58-.] | 0.124 |
| Cooked liver | 17 | 3 | 17.65 | 12 | 0 | 0 | . | [0.58-.] | 0.124 |
| Stew | 17 | 3 | 17.65 | 12 | 5 | 41.67 | 0.3 | [0.04-2.2] | 0.154 |
| Kisra (bread) | 17 | 3 | 17.65 | 12 | 5 | 41.67 | 0.3 | [0.04-2.2] | 0.154 |
| Kunafa (pastry dessert) | 17 | 7 | 41.18 | 12 | 8 | 66.67 | 0.35 | [0.06-2.04] | 0.176 |
| Lemonade | 17 | 4 | 23.53 | 12 | 1 | 8.33 | 3.38 | [0.27-181.5] | 0.286 |
| Mixed green salad | 17 | 7 | 41.18 | 12 | 7 | 58.33 | 0.5 | [0.09-2.8] | 0.362 |
| Other drink | 17 | 8 | 47.06 | 12 | 4 | 33.33 | 1.78 | [0.31-11.2] | 0.46 |
| Carrot salad | 17 | 3 | 17.65 | 12 | 1 | 8.33 | 2.36 | [0.16-134.8] | 0.474 |
| Spicy sauce | 17 | 9 | 52.94 | 12 | 5 | 41.67 | 1.58 | [0.28-9.1] | 0.55 |
| Pickled salad | 17 | 2 | 11.76 | 12 | 2 | 16.67 | 0.67 | [0.04-10.8] | 0.706 |
| Hummus | 17 | 2 | 11.76 | 12 | 2 | 16.67 | 0.67 | [0.04-10.8] | 0.706 |
| Mixed carrot salad | 17 | 2 | 11.76 | 12 | 1 | 8.33 | 1.47 | [0.07-94.3] | 0.765 |
| Basbousa (cake) | 17 | 6 | 35.29 | 12 | 4 | 33.33 | 1.09 | [0.18-7.1] | 0.913 |
| Aubergine salad | 17 | 4 | 23.53 | 12 | 3 | 25 | 0.92 | [0.12-7.9] | 0.927 |

Firth regression revealed that there was no dose response evident from the consumption of raw liver, (table 4), but it was evident for lamb; those eating a large portion were 77 times (95%CI 1.2-4848.8; p=0.04) more likely to be cases than non-cases. When examining dichotomous variables for lamb and raw liver consumption, lamb was not a significant risk factor but those eating raw liver were 28 times (1.4-546.9; p=0.03) more likely to be cases than non-cases. Stratification of the three foods revealed that the lamb and spicy sauce were not significant risk factors for illness and confounded the effect of consuming raw liver. Those eating raw liver were 50 times (1.7-1461.5; p=0.02) more likely to be cases than non-cases (table 5). Neither lamb nor spicy sauce improved the fit of our model but raw liver did (LR Chi^2^ = 7.93; p=0.005). We also tested for interaction with sex and age because barbecue attendees reported than males and females sat in separate groups and had differential access to food items. Children were mostly with their mothers. These factors yielded no significant interactions; sex and age did not adjust the likelihood of being a case.

**Table 4.** Firth univariate regression result for food consumed at lunch and barbecue.

| Food | Cases | Controls | OR | p-value | 95%CI |
| --- | --- | --- | --- | --- | --- |
| **Lunch** |  |  |  |  |  |
| Raw liver |  |  |  |  |  |
| small portion | 2 | 0 | 4.31 | 0.36 | [0.19-98.5] |
| Raw kidney |  |  |  |  |  |
| small portion | 2 | 0 | 4.31 | 0.36 | [0.19-98.1] |
| **Barbecue** |  |  |  |  |  |
| Raw liver |  |  |  |  |  |
| any portion | 9 | 0 | **27.94** | **0.03** | **[1.43-546.9]** |
| small portion | 3 | 0 | 10.29 | 0.14 | [0.47-225.9] |
| medium portion | 2 | 0 | 7.35 | 0.22 | [0.31-173.1] |
| large portion | 4 | 0 | 13.24 | 0.1 | [0.63-279.2] |
| BBQ lamb |  |  |  |  |  |
| any portion | 17 | 9 | 12.89 | 0.1 | [0.60-276.9] |
| small portion | 20 | 16 | 8.87 | 0.17 | [0.39-199.6] |
| medium portion | 3 | 2 | 9.8 | 0.19 | [0.33-287.4] |
| large portion | 5 | 0 | **77** | **0.04** | **[1.22-4848.8]** |
| Spicy sauce |  |  |  |  |  |
| any portion | 16 | 12 | 1.52 | 0.57 | [0.36-6.4] |
| small portion | 5 | 4 | 1.22 | 0.81 | [0.25-6.1] |
| medium portion | 0 | 1 | 0.33 | 0.52 | [0.01-9.6] |
| large portion | 4 | 0 | 9 | 0.16 | [0.41-198.2] |

**Table 5.** Stratified Firth multivariable regression results of suspected transmission vehicle food items consumed at barbecue.

| Food (barbecue) | OR | p-value | 95%CI |
| --- | --- | --- | --- |
| BBQ lamb | 6.87 | 0.24 | [0.28-165.9] |
| Raw liver | 49.99 | **0.02** | **[1.71-1461.5]** |
| Spicy sauce | 0.26 | 0.21 | [0.03-2.2] |

### Microbiology and WGS results

Microbiological results were available for ten individuals. Nine were PCR positive for Salmonella Spp., and five were cultured. All five were confirmed as *Salmonella Typhimurium*. Additionally, these five cases were identified as being genetically indistinguishable from one another (0 SNP difference) based on WGS undertaken by the GBRU. There were three cases with more than one pathogen in their stool sample (including two co-infections with Shiga-toxin producing *Escherichia Coli* (STEC) and one with *Campylobacter* Spp.)

### Food microbiology results

Environmental Health Officers retrieved 55 samples of leftover raw lamb, of which 11 were sent for culture testing and genomic sequencing at the GBRU. Samples for other food products used at the barbecue, including raw sheep’s liver, were unavailable. Sequences obtained from all 11 samples of raw lamb were genetically indistinguishable from one another, and those in human isolates from barbecue attendees (0 SNP). Identical genetic sequences to those in this outbreak’s cases were also returned from samples taken from the local butchers and supplying abattoir.

## Discussion

The traditional raw liver dish ‘marara’ was the primary vehicle of transmission in this outbreak. Microbiological evidence suggests that there was widespread contamination of sheep meat consumed at the barbecue, and that cross-contamination with other dishes occurred. However, consumption of infected raw sheep’s liver was likely to have resulted in a high infective dose and therefore an unusually high rate of severe illness. Many cases in this outbreak were severely unwell, with over half (54%, 12/22) of cases seeking emergency hospital care and 27% (6/22) being admitted overnight. Whilst we could not find a statistically significant association between raw liver consumption and hospital attendance, this may have been due to the small sample size. Two cases were also co-infected with STEC, both of whom were hospitalised due to their symptoms. The dual infection might explain the severity of their symptoms but field interview notes also revealed the high quantity of meat reported in their diets and also cross-contamination issues around food preparation practices.

It was not possible to establish where this contamination occurred in the food chain. Whilst genetically indistinguishable sequences were returned from human, meat and environmental samples, it is possible that the cooking of the lamb was enough to reduce the infective dose present in the meat at the barbecue, thus affecting people differently through quantity consumed and thoroughness of cooking.

The day of the barbecue was also very hot and there could have been an amplification effect due to the raw liver and lamb being unchilled for a long period. Our questionnaire responses also indicated that some of the lamb cooked on the barbecue was pink inside and that the marinade from the raw liver was used as a dip for other food and that one person used the plate the raw liver was served on as their plate to eat their meal from. All of this offers multiple opportunity for cross contamination. Another factor might be the preference of eating older animals in some cultures due to its flavour. This might be associated with a cumulative increase in pathogens in their organs (especially liver) and increased stress at slaughter, which exacerbates this phenomenon further.

### Public health messaging implications

Whilst there are many studies focusing on foodborne outbreaks of *Salmonella* infection, the majority of the outbreaks of salmonella which occur in the UK are associated with consumption of poultry and egg products [10]. This outbreak is one of few reported *Salmonella Typhimurium* outbreaks associated with consumption of lamb in the UK [11] [12]. Similarly, whilst dishes made from poultry liver, such as chicken and duck, are well known for being high risk foods for campylobacter across the UK and Europe [1] [2] there are very few studies describing outbreaks of salmonella linked to offal [15]. We identified one outbreak study from Australia, which identified an association between consumption of both raw and cooked lamb’s liver and *Salmonella* Typhimurium PT 197 in members of the Lebanese community [16]. Salmonella Typhimurium has been identified in samples taken from swabs of pig’s livers and tongue in pigs slaughtered for human offal consumption in Portugal, though were not linked to any human cases [17]. Frequently mararra also includes boiled sheep’s intestines, though this ingredient was not reported on this occasion.

This point source outbreak acted as a sentinel event, which through further case finding and sequencing led to the identification of samples genetically identical to the outbreak cases, in individuals who were resident of, or visitors to, Cardiff, who had not attended the barbeque or the lunch. Investigations are ongoing, following non-attendee cases also reporting consumption of lamb in Cardiff around the same time.

In addition to the 0-SNP cluster identified, in the five sequenced cases in this outbreak were identified as being part of a larger ongoing 5 SNP level cluster; 1.26.136.634.2037.4574. %. Cases in this cluster date back to 2018 and are currently spread across Wales, with cases also in Scotland, London and the South West and Midlands of England. At least two of these cases in this wider cluster were in people with occupational contact with sheep farms, suggesting the possibility of a transmission chain with a farm origin. Cases in this cluster have continued to be identified following the close of the barbeque investigation, and investigations continue into the source of these infections and food supply chains.

## Community Engagement

Eid al-Adha (‘Festival of Sacrifice’) is one of the most important festivals in the Muslim calendar. The festival remembers the prophet Ibrahim’s willingness to sacrifice his son when God ordered him to. In some countries, Muslims sacrifice a sheep or goat (in Britain the animal is killed at a slaughter house) and the meat is shared equally between family, friends and the poor. Eid usually starts with Muslims going to the Mosque for prayer and is a time when they visit family and friends and also give money to charity.

As part of the outbreak response, Public Health Wales sought engagement advice from experts in its service user experience team. Options were considered for engaging the North African community in Cardiff, but also the various communities in Wales, on food safety whilst remaining culturally sensitive and maintaining anonymity. With the support of an engagement coordinator from Cardiff council, an educational session was conducted during the Eid-al-Adha festival which included members of the community that consume the traditional “marrara” dish and other communities that celebrate Eid al-Adha. Key food safety and hygiene messages were presented followed by an open conversation. The session received positive verbal feedback from the participants. Learning from this engagement helped Public Health Wales develop a universal response (box 1), available in several languages, to food safety that is now programmed into annual communications for World Food Safety Day in June and ahead of specific festivals and events such as Eid al-Adha. The approach consisted of two phases:

**Phase one**: general food safety messages shared via social media on World Food Safety Day in June 2022, available in several languages (figure 3).

**Figure 2.**
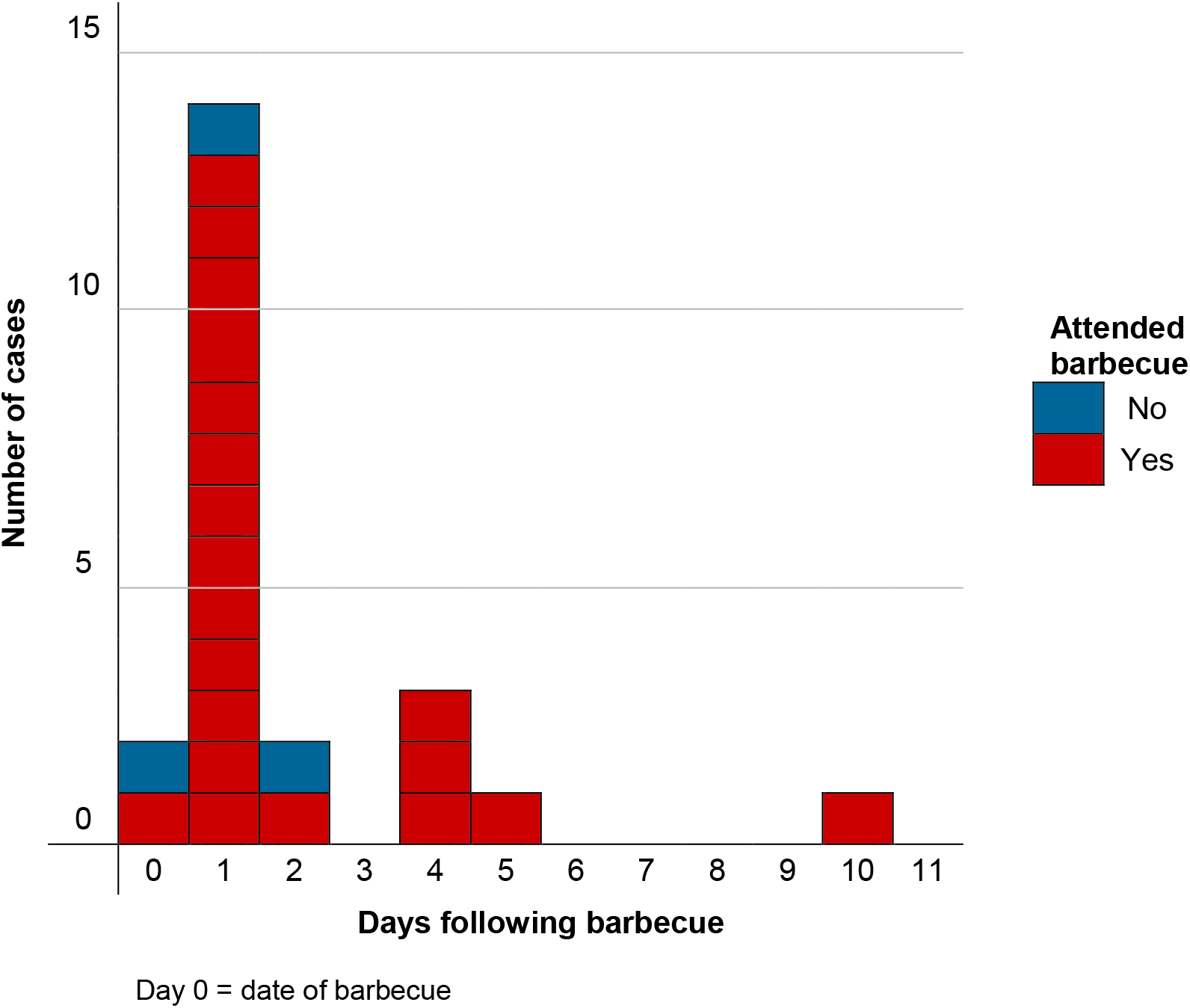
Epidemic curve: Day of symptom onset in confirmed cases (n=22) by barbecue attendance.

**Figure 3.**
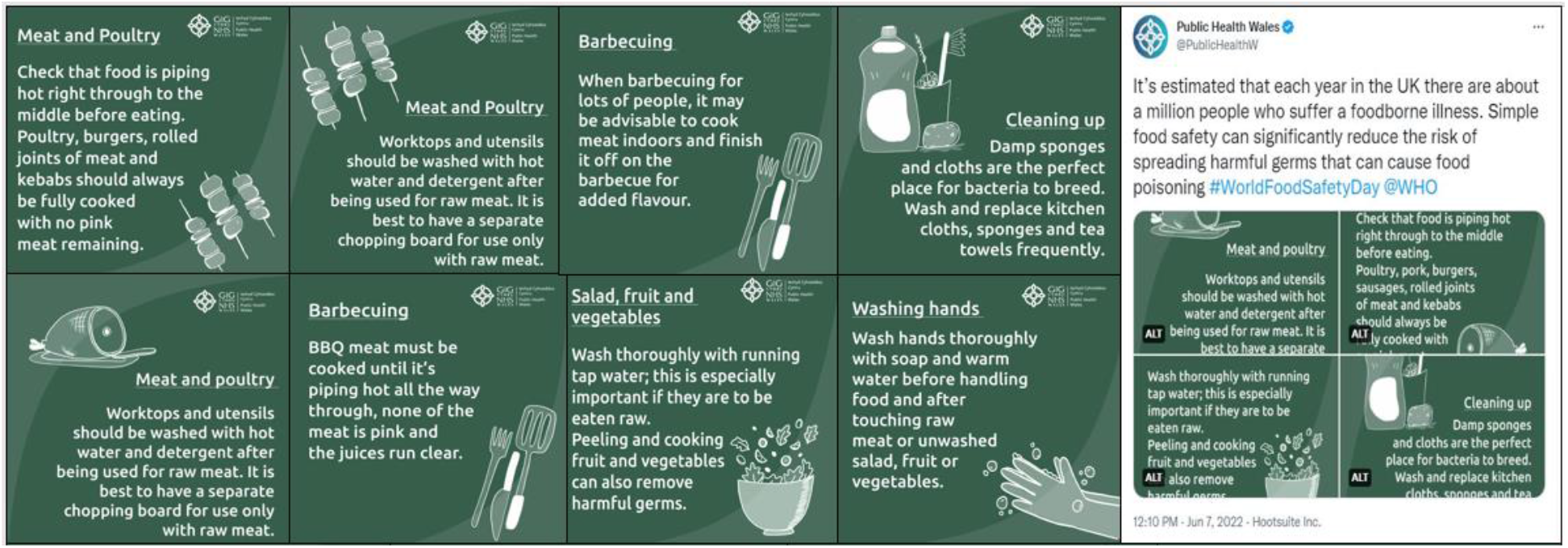
Public health messaging developed through community engagement and feedback as part of the outbreak.

Phase two: Food safety infographics tailored for the Eid al Adha festival in July 2022. Infographics were shared via Public Health Wales’s social media platforms, external partners and the WhatsApp group established for sharing pandemic information with communities.

### Box 1.

***Lessons learnt from community engagement:***

1. Setting clear and realistic engagement objectives from the project outset
2. Ensure resources are adequate to deliver on objectives
3. Allow enough time for translations of materials.
4. Ensure partnership working with other statutory partners and align the engagement where possible
5. Establishing early contact with communities and partners in plenty of time to ensure maximum publicity and participation
6. Ensure networks are maintained regularly so there is willingness to help when you need support in the future

## Conclusions

Whilst the consumption of raw or undercooked meats is associated with salmonellosis outbreaks in Europe [6] [7], this outbreak in a minority ethnic community in South Wales identified a new vehicle of interest, a traditional raw offal dish. Future outbreak investigations, particularly where cultural events are associated with particular foods, should consider dishes beyond those on routine questionnaires to those which may be relevant to the community in question.

Consumption of raw and undercooked meat and offal continues to be an ongoing public health risk. Improving awareness and proactive messaging, especially to communities who consume raw/undercooked meat or offal either regularly, or during specific religious or cultural events, may help reduce this risk.

## Acknowledgements

We wish to thank the IMT for their work on the outbreak and on the subsequent wider investigation of linked cases. We are grateful for the Public Health Wales health protection team and the local environmental health team for their contribution to the investigation of the outbreak, in particular Rachel Andrew and Su Mably. We would like to thank Katie Allen from the Public Health Wales Communications Teams for their advice on and assistance on producing specific food safety resources for community engagement. To Anais Painset from UKHSA’s Gastro-bacterial Reference Unit (GBRU) for the microbiological analysis of human and food samples associated with the incidents described in the paper and subsequent whole genome sequencing; and to the APHA for their investigations into the food chain that the meat followed.

## Authors’ contributions

DT designed and led the epidemiological investigation. JA, CS, GH and DT drafted the manuscript. JI, IM and GK provided information on the steps of community engagement. CS, GH, EC, JA, CW, LF, OO, AP, RS, AJ and DT carried out epidemiological investigations and undertook interviews with the cohort, complied the descriptive analysis and contributed to the epidemiological investigation. HH led the microbiological investigation. All authors contributed to the writing and revision of this manuscript and approved the final version.

## Ethics

Ethical oversight of the project was provided by the PHW Research and Development Division. As this work was carried out as part of the health protection response to a public health emergency in Wales, using routinely collected surveillance data, PHW Research and Development Division advised that NHS research ethics approval was not required. The use of named patient data in the investigation of communicable disease outbreaks and surveillance of notifiable disease is permitted under PHW’s Establishment Order. Data were held and processed under PHW’s information governance arrangements, in compliance with the Data Protection Act, Caldicott Principles and PHW guidance on the release of small numbers. No data identifying protected characteristics of an individual were released outside of the IMT.

## Data Availability Statement

The data used in this investigation contains personal identifiable information. Anonymised information, including that contained in the supplementary information, required to reproduce these results is available from the corresponding author on reasonable request.

## References

1. Hawker J, et al. (2005) Communicable Disease Control and Health Protection Handbook, 2nd edn. Oxford: Blackwell, pp. 193.

2. UK Health Security Agency. (2020) UK SMI ID 24: identification of Salmonella species. Published online December 2020: https://assets.publishing.service.gov.uk/government/uploads/system/uploads/attachment_data/file/936585/ID_24_do+.pdf.

3. Public Health England. (2019) Routine reports of gastrointestinal infections in humans, England and Wales: September and October 2019.” London, 2019. Published online November 2019: https://assets.publishing.service.gov.uk/government/uploads/system/uploads/attachment_data/file/845643/hpr4019_GIs.pdf.

4. Silva C, Calva E, Maloy S. (2014) One Health and Food-Borne Disease: Salmonella Transmission between Humans, Animals, and Plants. Journal of Microbiology Spectrum. 2:, ePub DOI: 10.1128/microbiolspec.oh-0020-2013.

5. Public Health Wales. (2022) Laboratory reports of SALMONELLA in Wales 2018-2021,” Published online January 2021: https://public.tableau.com/app/profile/public.health.wales.health.protection/viz/LaboratoryreportsofSALMONELLAinWales2014-2017/Dashboard2.

6. Ashton P M, et al. (2015) Whole genome sequencing for the retrospective investigation of an outbreak of Salmonella Typhimurium DT 8. PLoS Currents OUTBREAKS. Published online February 2015: 10.1371/currents.outbreaks.2c05a47d292f376afc5a6fcdd8a7a3b6.

7. Animal and Plant Health Agency. (2021) Salmonella in Livestock Production in Great Britain. Published online September 2022: https://assets.publishing.service.gov.uk/government/uploads/system/uploads/attachment_data/file/1060634/salm-livestock-prod-gb20.pdf.

8. SmartSurvey. SmartSurvey overview (https://www.smartsurvey.co.uk/survey-software.) Accessed 16 October 2021

9. Language Line. Language Line services (https://www.languageline.com/uk.)Accessed August 2022)

10. Public Health England. (2021) Non-typhoidal Salmonella data 2010 to 2019. Published online October 2021: https://assets.publishing.service.gov.uk/government/uploads/system/uploads/attachment_data/file/1026208/salmonella-annual-report-2019.pdf

11. Evans MR, et al. (1999) An outbreak of Salmonella typhimurium DT170 associated with kebab meat and yoghurt relish. Journal of Epidemiology and Infection. 122: 377–383. DOI: 10.1017/S0950268899002253.

12. Food Safety News. (2018) Almost 300 sick and one dead due to Salmonella in UK. Published online October 2018: https://www.foodsafetynews.com/2018/10/almost-300-sick-and-one-dead-due-to-salmonella-in-uk/#:~:text=Authorities%20in%20England%20and%20Scotland,had%20been%20detected%20in%20England.

13. Little CL, et al. (2010) A recipe for disaster: Outbreaks of campylobacteriosis associated with poultry liver pâté in England and Wales. Journal of Epidemiology and Infection. 138: 1691–1694. DOI: 10.1017/S0950268810001974.

14. Bintsis T, (2017) Foodborne pathogens. AIMS Microbiology. 3: 529–563. DOI: 10.3934/microbiol.2017.3.529.

15. Cornell J, Neal KR. (1998) Protracted outbreak of Salmonella typhimurium definitive phage type 170 food poisoning related to tripe, ‘pig bag’, and chitterlings. Journal of Communicable Disease and Public Health 1: 28–30. DOI: https://pubmed.ncbi.nlm.nih.gov/9718834/

16. Heiss J, et al. (2008) A Salmonella Typhimurium 197 outbreak linked to the consumption of lambs’ liver in Sydney, NSW. Journal of Epidemiology and Infection. 136: 461–467. DOI: 10.1017/S0950268807008813.

17. Vieira-Pinto M, et al. (2018) Salmonella sp. in edible offal (liver and tongue) from pigs slaughtered for consumption. In: International Conference on the Epidemiology and Control of Biological, Chemical and Physical Hazards in Pigs and Pork,” pp. 202–205. DOI: 10.31274/safepork-180809-621.

18. Ethelberg S, et al. (2007) Outbreak with multi-resistant Salmonella Typhimurium DT104 linked to carpaccio, Denmark, 2005. Journal of Epidemiology and Infection 135: 900–907. DOI: 10.1017/S0950268807008047.

19. Brandwagt D et al. (2018) Outbreak of Salmonella Bovismorbificans associated with the consumption of uncooked ham products, the Netherlands, 2016 to 2017. Eurosurveillance 23: 1–6. DOI: 10.2807/1560-7917.ES.2018.23.1.17-00335.

